# Intensified Blood Pressure Control During Hospital Admission and on Discharge: A Systematic Review and Meta-Analysis

**DOI:** 10.1101/2025.04.22.25325620

**Authors:** Yevhen Kushnir, Nelson Barrera, Pedro Arias-Sanchez, Erick Romero, Iurii Statnii, Anton Stolear, Kristina Golovataya, Maria Fernanda Solorzano, Evgeny Shkolnik

**Author notes:** co-first author. **Address for correspondence: Yevhen Kushnir**, MD, Nelson **Barrera**, MD **SBH Health System**, Department of Internal Medicine, NY **10457**, **United States;**, **Evgeny Shkolnik**, MD, FACC; Yale University School of Medicine, **267 Grant St**, **Bridgeport**, CT **06610;**. **Funding:** This research has not been funded directly by any organization. **Disclosure statement:** The remaining authors have nothing to disclose.

## Abstract

**Background:** Limited single-center studies suggest that intense blood pressure (BP) control during non-cardiac admissions with elevated BP may lead to worse outcomes.

**Objectives:** We aimed to perform a systematic review and meta-analysis exploring the safety of intensified BP control compared to a more conservative approach in patients with asymptomatic elevated BP in non-cardiac admissions and on discharge.

**Methods:** We searched PubMed, Cochrane, and Embase for studies comparing intensified vs. conservative management of elevated BP during hospital stay in patients with non-cardiac admissions. The primary outcomes were stroke, acute kidney injury (AKI), myocardial infarction (MI), and length of stay (LOS).

**Results:** Four studies of 77448 patients were included, of whom 38724 underwent intensified BP control vs 38724 conservative. The follow-up period ranged from 1 to 365 days. The pooled analysis showed a significant difference in stroke (OR 3.69; 95% CI 1.47-9.28; p <0.006), AKI (OR 1.22; 95% CI 1.14-1.30; p<0.00001), longer LOS (MD 1.52; 95% CI 1.11-1.93 p<0.00001) and nonsignificant difference in MI (OR 2.08; 95% CI, 0.84-5.16, p = 0.12) in the intensified BP management group.

**Conclusion:** This meta-analysis suggests that an intensified control of asymptomatic elevated BP during non-cardiac hospitalization and on discharge is associated with higher odds of stroke, AKI, and prolonged length of stay when compared with conservative control.

**CONDENSED ABSTRACT:** Studies suggest that intense blood pressure (BP) control during non-cardiac admissions with elevated BP may lead to worse outcomes. We performed a systematic review and meta-analysis of 77448 patients from 4 non-randomized studies exploring the safety of intensified BP control compared to a more conservative approach in patients with asymptomatic elevated BP in non-cardiac admissions and on discharge. Our results suggest that intensified blood pressure control was associated with higher odds of stroke, AKI, and prolonged length of stay when compared with conservative control. Further randomized controlled trials are need it to support our results.

## Introduction

Hypertension affects approximately 50% of individuals aged over 20 years and remains a significant risk factor for cardiovascular diseases, stroke, and mortality. ^1^ ^2^ Among hospitalized patients, the prevalence of hypertension is particularly high, ranging from 50.5% to 72%.^3^ Despite this, current clinical guidelines provide limited guidance on the intensification of antihypertensive treatment during hospitalization and on discharge, leaving a gap in evidence-based recommendations for managing this condition in acute care settings.

The intensification of antihypertensive treatment during hospitalization has been inconsistent, resulting in 37% to 77% of hypertensive patients remaining hypertensive at the time of discharge.^3^ Furthermore, many patients with inpatient hypertension continue to exhibit elevated blood pressure levels during outpatient follow-up^3^, underscoring the need for improved management strategies. However, changes in medication regimens may put patients at higher risk of adverse reactions.^4^ A study in an emergency department found that inpatient blood pressure readings can differ significantly from the patient’s home blood pressure level.^5^ Moreover, patients discharged with substantially escalated antihypertensive regimens have been reported to experience hypotension during follow-up visits.^5^

The 2019 American Heart Association guideline do not recommend a specific approach for asymptomatic inpatients with elevated blood pressure.^6^ However, subsequent observational studies suggest that more intensive blood pressure control may be associated with adverse outcomes;^3,7^ The 2024 American Heart Association statement consolidates findings from previous studies and advocates that treatment for asymptomatic elevated blood pressure should generally be the exception rather than the rule, noting the need for further studies to clarify whether clinical benefits exist for asymptomatic patients with markedly elevated blood pressure.^8^ In light of the ongoing lack of consensus regarding the risks and benefits of blood pressure control in asymptomatic patients, this study aims to offer an updated systematic review and meta-analysis of the characteristics and outcomes related to inpatient hypertension management and follow-up at discharge.

## Methods

This systematic review and meta-analysis were performed and reported following with the Cochrane Collaboration Handbook for Systematic Reviews of Interventions and the Preferred Reporting Items for Systematic Reviews and Meta-Analysis (PRISMA) Statement guide.^9^ The prospective meta-analysis protocol was registered at the International Prospective Register of Systematic Reviews (PROSPERO; <u>#CRD42024566609</u>.) in July 2024.

### Data source and search strategy

We systematically searched PubMed, Embase, and Cochrane from inception to January 2025 using the terms “inpatient blood pressure control outcomes.” Additionally, we manually searched the references from all included studies for additional studies. Two authors (Y.K and N.B.) independently screened titles and abstracts and evaluated the articles in full for eligibility based on prespecified criteria. Discrepancies were resolved in a panel discussion between authors.

### Eligibility Criteria

We considered studies eligible for inclusion if they: (1) were randomized controlled trials (RCTs) or non-randomized cohorts, (2) comparison of intensified versus non-intensified elevated blood pressure control during hospital admissions, (3) patients with non-cardiac admissions, and (4) follow-up duration of more than one day.

We defined the “intensified blood pressure control group” as a new or higher dose or IV or PRN or PRN plus scheduled anti-hypertensive medication in the regimen. We defined the “non-intensified blood pressure control group” as those before admission or scheduled or no PRN anti-hypertensive medication in the regimen. We defined “non-cardiac admissions” as conditions that do not require specific blood pressure control goals and regimens (patients with a hypertensive emergency, stroke, MI, or aortic dissection were excluded).

The follow-up duration was select to assess the intervention’s short- and long-term outcomes. Studies were excluded if they (1) lacked a control group or (2) involved a cardiac cause of admission. Definitions of intensified blood pressure control varied slightly across studies and are detailed in Table 1.

**Table 1.**
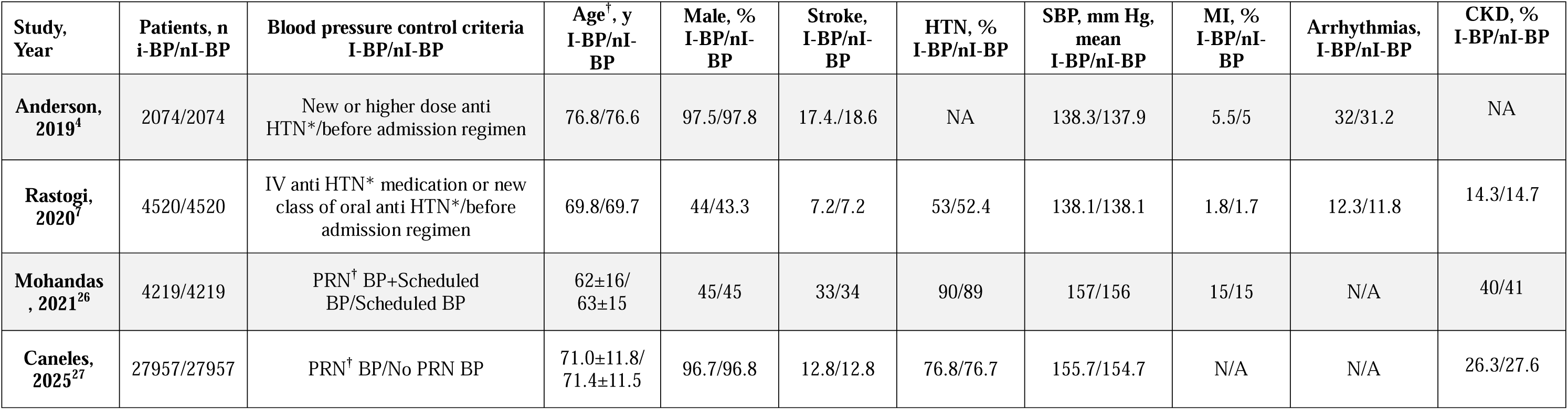
Baseline characteristics of the included studies. Abbreviations: I-BP: intensified blood pressure group; nI-BP: non-intensified blood pressure group; SBP – systolic blood pressure; MI – myocardial infarction; HTN-hypertension, CKD – chronic kidney disease Notes: *-antihypertensive medication; **^†^** - as needed

| Study, Year | Patients, n<br>i-BP/nI-BP | Blood pressure control criteria<br>I-BP/nI-BP | Age<sup>†</sup>, y<br>I-BP/nI-BP | Male, %<br>I-BP/nI-BP | Stroke,<br>I-BP/nI-BP | HTN, %<br>I-BP/nI-BP | SBP, mm Hg,<br>mean<br>I-BP/nI-BP | MI, %<br>I-BP/nI-BP | Arrhythmias,<br>I-BP/nI-BP | CKD, %<br>I-BP/nI-BP |
| --- | --- | --- | --- | --- | --- | --- | --- | --- | --- | --- |
| **Anderson, 2019<sup>4</sup>** | 2074/2074 | New or higher dose anti HTN*/before admission regimen | 76.8/76.6 | 97.5/97.8 | 17.4/18.6 | NA | 138.3/137.9 | 5.5/5 | 32/31.2 | NA |
| **Rastogi, 2020<sup>7</sup>** | 4520/4520 | IV anti HTN* medication or new class of oral anti HTN*/before admission regimen | 69.8/69.7 | 44/43.3 | 7.2/7.2 | 53/52.4 | 138.1/138.1 | 1.8/1.7 | 12.3/11.8 | 14.3/14.7 |
| **Mohandas, 2021<sup>26</sup>** | 4219/4219 | PRN<sup>†</sup> BP+Scheduled BP/Scheduled BP | 62±16/<br>63±15 | 45/45 | 33/34 | 90/89 | 157/156 | 15/15 | N/A | 40/41 |
| **Caneles, 2025<sup>27</sup>** | 27957/27957 | PRN<sup>†</sup> BP/No PRN BP | 71.0±11.8/<br>71.4±11.5 | 96.7/96.8 | 12.8/12.8 | 76.8/76.7 | 155.7/154.7 | N/A | N/A | 26.3/27.6 |

### Endpoints

The outcomes of interest were acute kidney injury (AKI), stroke, myocardial infarction (MI), and length of stay (LOS).

### Quality assessment

Non-randomized studies were assessed using the Risk of Bias in Non-randomized Studies of Interventions tool (ROBINS-I).^10^ Two independent authors (Y.K. and N.B.) conducted the risk of bias assessment. Disagreements were resolved through consensus after discussing the reasons for the discrepancies. Publication bias was evaluated using funnel-plot analysis of point estimates about study size.

### Statistical analysis

Odds ratios (OR) with 95% confidence intervals (CI) were used to compare treatment effects for categorical endpoints. Continuous outcomes were compared using standardized mean differences (MD). Heterogeneity was assessed using I^2^ statistics and the Cochran Q test; p values <0.10 and I² >25% were considered indicative of significant heterogeneity. We applied the DerSimonian and Laird random-effects models. Statistical analysis was performed using ReviewManager 5.4 (Cochrane Center, The Cochrane Collaboration, Denmark).

## Results

### 3.1 Study selection and baseline characteristics

Our systematic search yielded 1270 results. After removing duplicate records and ineligible studies, 12 studies remained and were fully reviewed based on the inclusion criteria; Of these, four non-randomized studies, comprising 77448 patients were included. Comprehensive details of the study selection are detailed in Figure 1.

**Figure 1.**
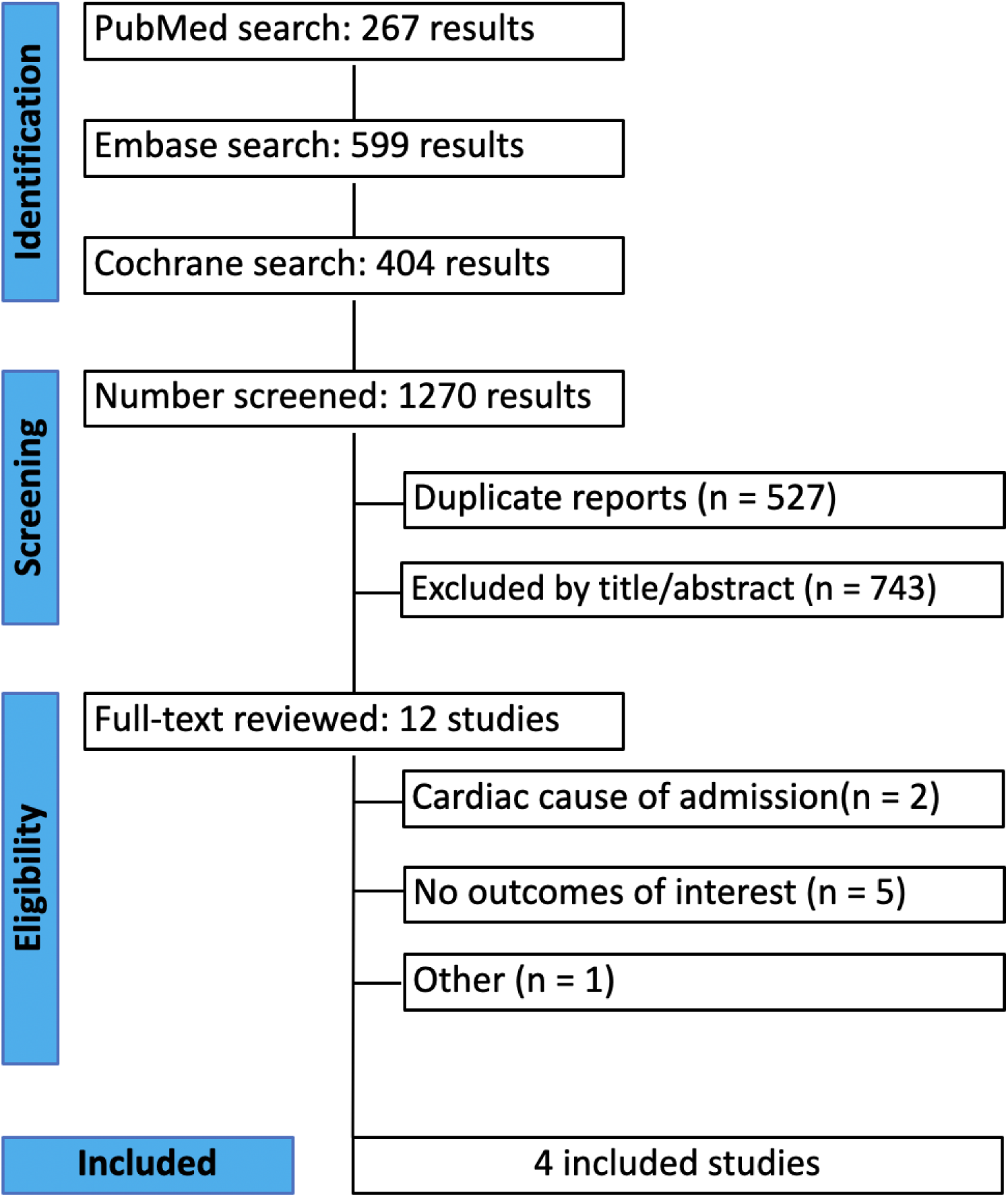
PRISMA flow diagram of study screening and selection.

A total of 38724 patients received an intensified blood pressure control regimen, while 38724 patients received a non-intensified blood pressure control regimen. The mean systolic blood pressure in the intensified group was 147.3 mm Hg, compared to 146.7 mm Hg in the non-intensified group. The mean age was 69.9 years in the intensified management arm and 70.1 years in the conservative arm. In the intensified group, 70.8% of patients were male, while 70.7% were male in the non-intensified group. The follow-up period ranged from 1 to 365 days. Table 1 summarizes the main characteristics of the included studies.

### 3.2 Pooled analysis of all studies

#### Incidence of stroke

The pooled analysis showed a significantly higher chance of stroke in the intensified blood pressure control group compared to the non-intensified control group (OR 3.69; 95% CI 1.47-9.28; p <0.006; Figure 2). Heterogeneity was moderate, with I² = 57%.

**Figure 2.**
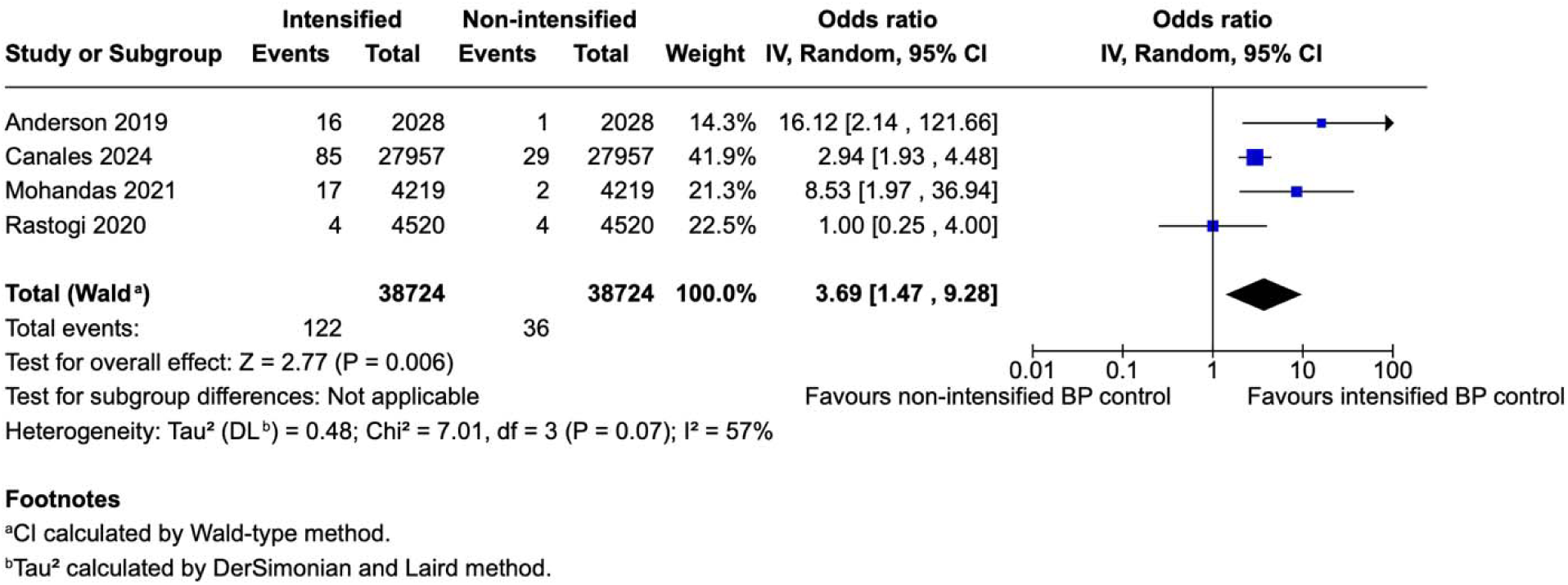
Forest plot showing the chance of stroke in intensified and non-intensified blood pressure control groups.

#### Incidence of AKI

The pooled analysis showed a significantly higher chance of AKI in the intensified blood pressure control group compared to the non-intensified control group (OR 1.22; 95% CI 1.14 – 1.30; p<0.00001; Figure 3). Heterogeneity was low, with I² = 24%.

**Figure 3.**
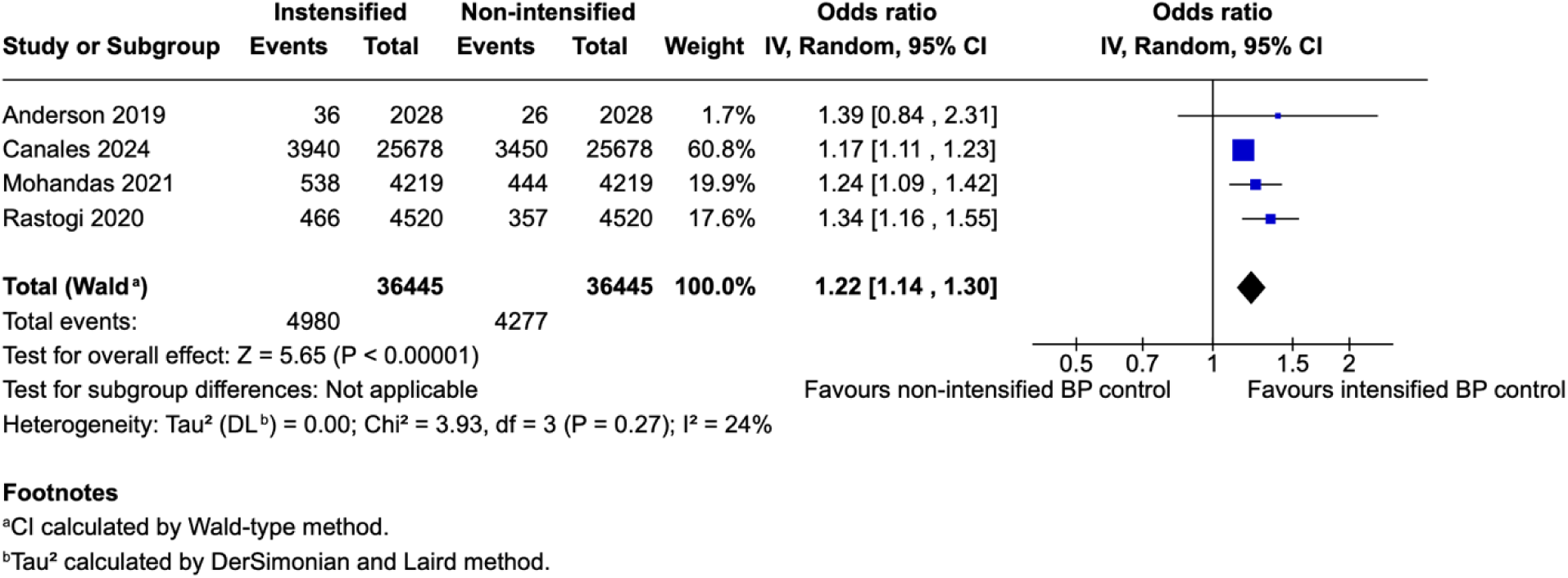
Forest plot showing the chance of AKI in intensified and non-intensified blood pressure control groups.

#### Length of stay

Intensified blood pressure control was associated with a significantly longer length of stay compared to non-intensified blood pressure control (MD 1.52; 95% CI 1.11-1.93; p<0.01; Figure 4). The results showed a high heterogeneity with I^2^ = 98%.

**Figure 4.**
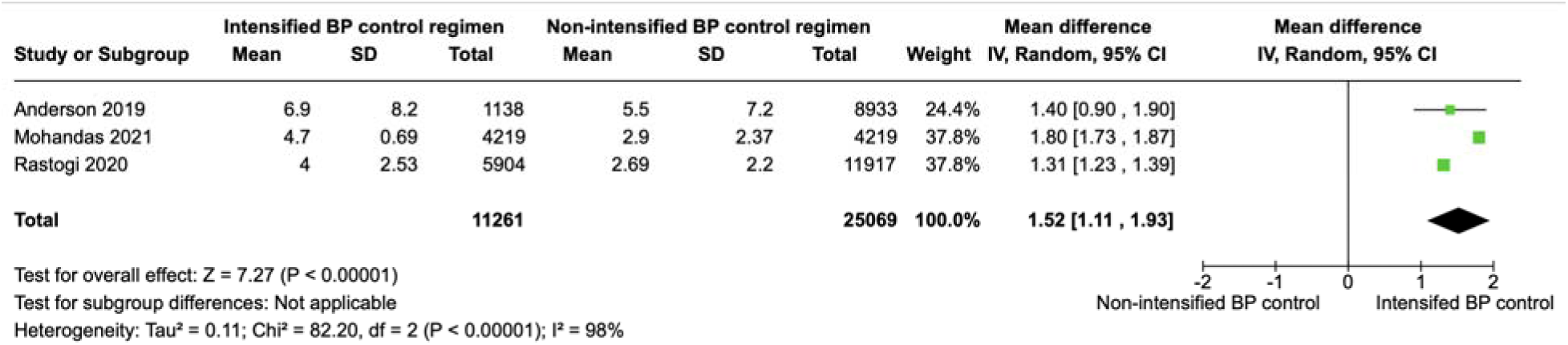
Forest plot showing the length of stay in intensified and non-intensified blood pressure control groups.

#### Myocardial infarction

There was no significant difference in the chance of myocardial infarction between the intensified blood pressure control group and the non-intensified blood pressure control group (OR 2.08; 95% CI 0.84-5.16; p<0.00001; Figure 5). The results showed a high heterogeneity with I^2^ = 94%.

**Figure 5.**
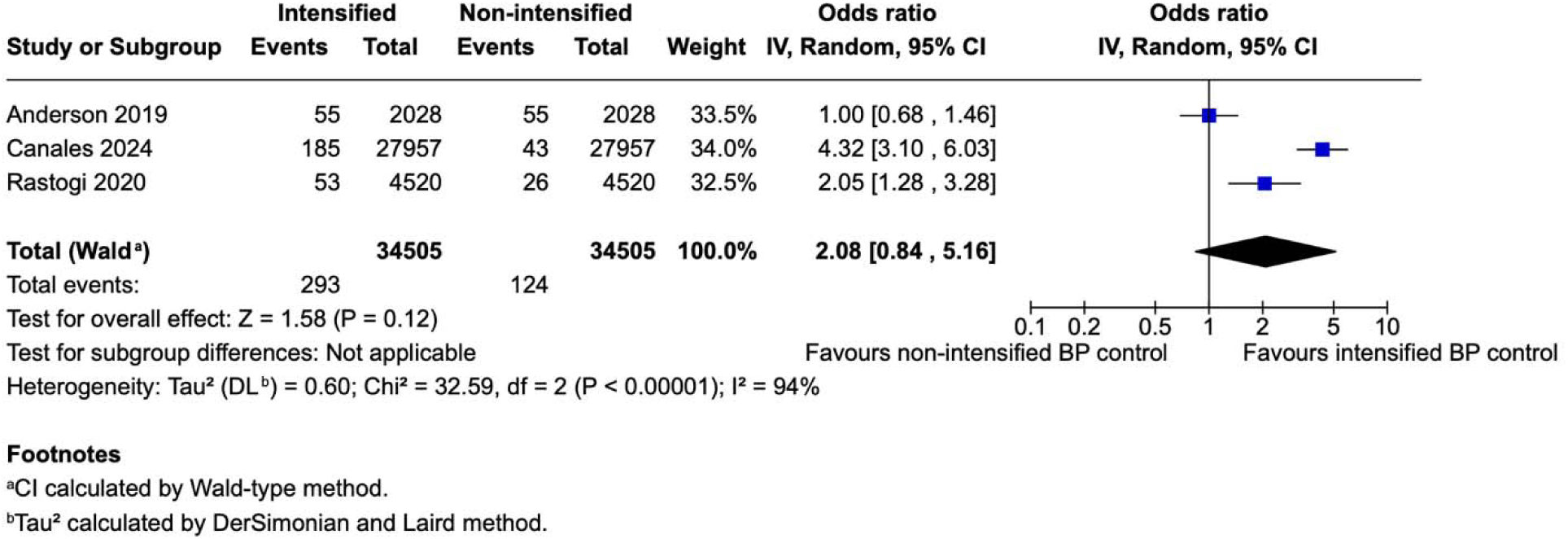
Forest plot showing the chance of myocardial infarction in intensified and non-intensified blood pressure control group.

#### Leave-one-out sensitivity analysis

Leave-one-out sensitivity analysis was performed to explore the impact of separate studies on the cumulative analysis for stroke and AKI. Overall, the results remained similar to the primary analysis. Sensitivity analysis results are presented in Figure 6 and Figure 7.

**Figure 6.**
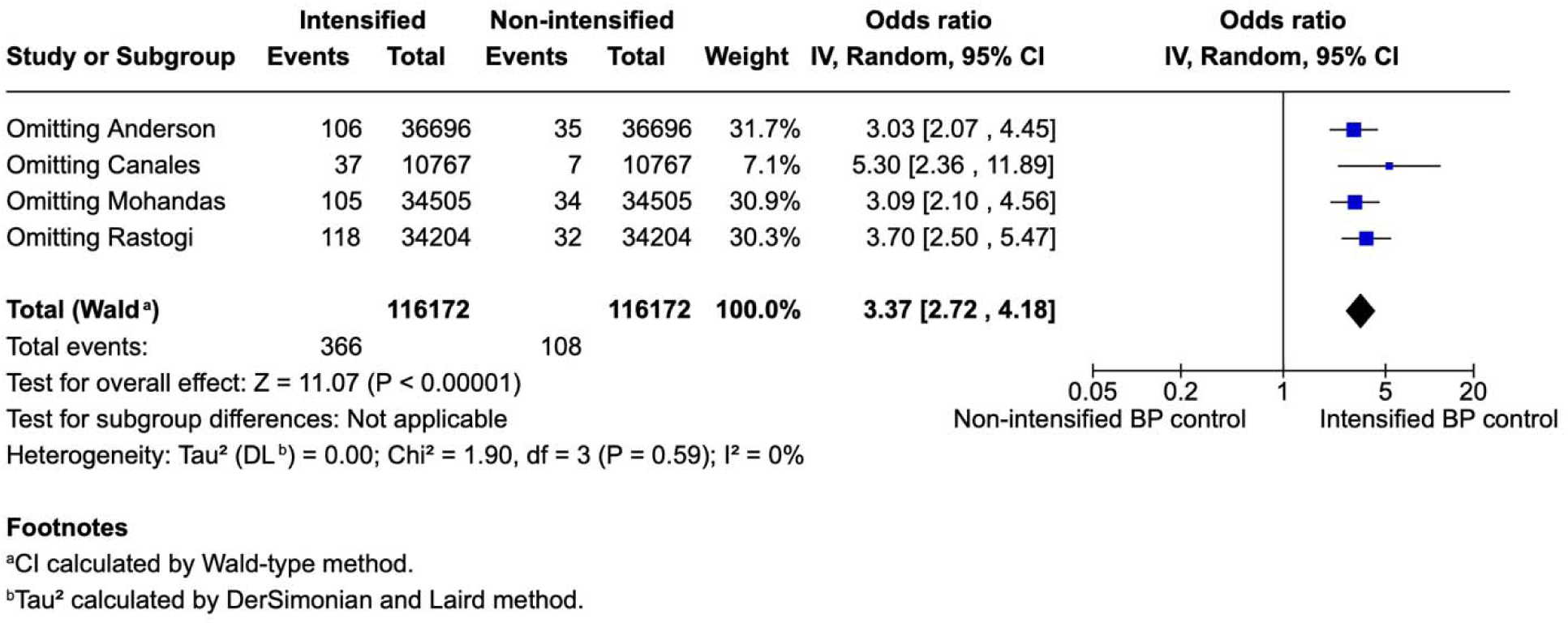
Forest plot showing leave-one-out sensitivity analysis for stroke.

**Figure 7.**
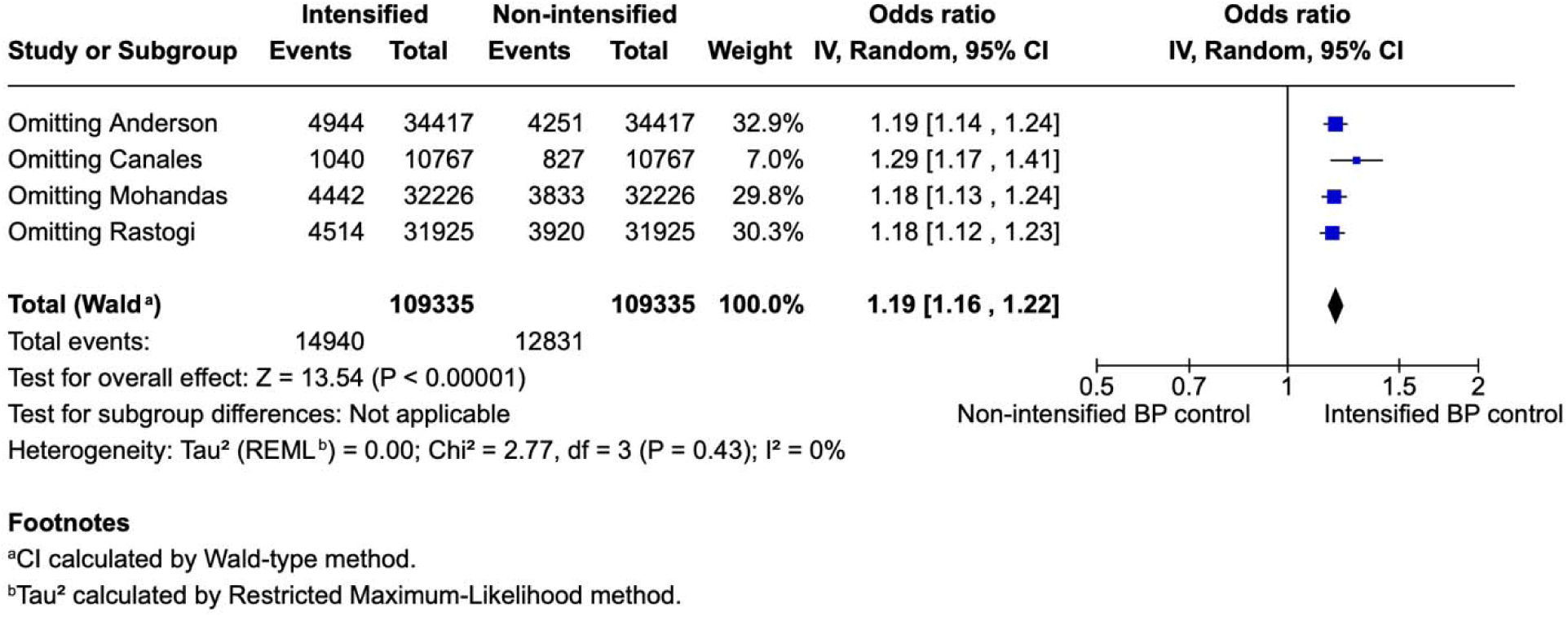
Forest plot showing leave-one-out sensitivity analysis for AKI.

#### Quality assessment

No studies were considered at high risk of bias as described in Supplementary Table 1 in supplementary appendix. In the funnel plot, studies occupied symmetrical distribution according to weight and converged toward the pool effect as the weight increase.

## Discussion

In this systematic review and meta-analysis, encompassing four studies and a total of 77448 patients, we compared the outcomes of intensified blood pressure control versus non-intensified blood pressure control in asymptomatic patients with elevated blood pressure, both at hospital admission and discharge.

1. Our findings revealed that the intensified blood pressure control group was associated with a 3.69 increase in the likelihood of stroke, a 1.22 increase in the odds of acute kidney injury (AKI), and a 1.52 longer length of hospital stay, compared to the non-intensified control group.
2. Although the odds of myocardial infarction (MI) were higher in the intensified control group, this result did not reach statistical significance.

This study builds upon previous research by providing a contemporary analysis of outcomes in intensified vs. non-intensified blood pressure patients using the highest-quality evidence available, addressing the limitations in earlier studies.

Asymptomatic elevated blood pressure (BP) was present in 50%-72% of hospitalizations.^3^ In patients with hypertensive emergency, the approach to blood pressure control is well established; a majority of hospital encounters with elevated blood pressure do not have any indication to pursue tight blood pressure control. The 2019 American Heart Association guideline does not recommend a specific approach due to the lack of evidence;^6^ however, the 2024 AHA statement points out that further studies are needed to clarify whether there is a clinical benefit for asymptomatic patients with markedly elevated blood pressure.^8^

Elevated BP readings can be a concerning finding during a hospital stay. A survey of 181 residents across three U.S. hospitals revealed that 44% would initiate or adjust antihypertensive treatment if the systolic blood pressure (SBP) ranged from 140 to 159 mmHg.^11^ Various factors drive the administration of PRN antihypertensive medication, including multiple cardiovascular risk factors, vital sign alarms, and nursing notifications.^8^ This approach can lead to additional reductions in BP and the subsequent withholding of scheduled antihypertensive medication. Consequently, this pattern may result in higher BP readings due to missed scheduled antihypertensive medication, prompting further PRN medication use and contributing to significant BP variability.

The hospital environment is teeming with numerous reversible causes for elevated BP. Key contributors include anxiety, sleep deprivation, psychological stress, and pain, all of which need to be carefully assessed and managed. In addition, various inpatient medications—such as IV fluids, corticosteroids, and NSAIDs—can also elevate BP. Moreover, inaccurate home medication reconciliation can lead to rebound hypertension. A retrospective review found that 41% of patients prescribed PRN antihypertensive medications did not receive their usual home BP regimen while hospitalized. Addressing these factors is crucial for effective BP management and preventing unnecessary complications.^12^

### Mechanisms

Autoregulation is a mechanism that enables organs such as the brain, heart, and kidneys to maintain stable blood perfusion by adjusting their vasculature resistance in response to fluctuations in perfusion pressure;^13^ in individuals with normal blood pressure, the brain’s vascular resistance adjusts over a wide range of mean arterial pressures to maintain consistent blood flow.^14^ However, in patients with chronically elevated blood pressure, the autoregulatory curve to the right, with a subsequent risk of hypoperfusion when administering antihypertensive medication, as the precise lower limit of autoregulation in individual hypertensive patients is often unknown.^14^ This shift in autoregulation could help explain the study’s findings, where intensified blood pressure control was associated with a significantly higher risk of stroke, AKI, and prolonged hospital stay.

### Stroke

Elevated BP is a well-established and highly impactful modifiable risk factor for stroke.^15^ The mean SBP in the intensified group was 147.3, and 146.7-mm Hg in the non-intensified group. It is challenging to explain the burden of adverse outcomes in heterogeneous populations with such a slight difference in mean SBP; adding the nature of autoregulation curve dynamics, we hypothesized that a level of blood pressure is not a single contributor to adverse effects but also a blood pressure variability, which a mean SBP values cannot fully reflect. Aggressive BP control can lead to substantial BP fluctuations, which might be more harmful than consistently elevated BP. ^16^ Higher BP fluctuations after stroke were associated with a higher risk of death within 90 days.^17^ Paradoxically, while efforts to control BP are aimed at preventing adverse outcomes, including stroke, such intensive management may inadvertently increase the risk of stroke.

Crucial data, such as atrial fibrillation was reported only in 1 study and their corresponding CHADS-VASc scores was not reported in any of the studies, which may lead to variations in the stroke risk between cohorts. The use of multiple blood pressure medications and different routes of administration, along with variable definitions of “intensified” and “non-intensified” groups and their patients, along with the lack of randomized trials, appear to significantly contribute to the moderate heterogeneity of results, which was corroborated in our sensitivity analysis.

### AKI

AKI is a common clinical event that affects up to 15% of hospitalized patients.^18^ 20–50% of AKI patients develop progressive CKD, while 3–15% reach end-stage kidney disease, all associated with increased mortality.^19^ Hypertension and hypotension are well-known risk factors for AKI; however, BP variability may also contribute. This mechanism can partially explain our study’s 1.22 increase in odds for AKI. In a study of 82659 patients with intraoperative arterial blood pressure monitoring during non-cardiac surgeries, higher intraoperative BP variability is associated with higher risks of postoperative AKI independent of hypotension and other clinical characteristics.^20^ Another study in patients undergoing surgery for acute aortic dissection showed that patients with AKI had higher SBP variability.^21^ Underlying chronic kidney disease(CKD) is a known risk factor for AKI;^22^ however, it is unlikely to account for a higher AKI rate in the intensified BP control group due to similar rate of CKD in both groups. One included study had a large portion of patients admitted with UTI, which is another risk factor for AKI.^4,23^ However, both groups had the same rates of UTI, and the removal of this study during sensitivity analysis did not affect the rate of AKI in the intensified group in pooled analysis.

### Myocardial infarction

A cohort study of 1.3 million admissions revealed an MI incidence of 4.27 per 1000 admissions.^24^ Due to the nature of the study population (noncardiac admissions) in our analysis and the low incidence of in-hospital myocardial infarction, lower events were expected. However, results may vary in a population with more cardiovascular risk factors. One study of 1828 intracranial hemorrhage survivors showed that higher BP variability was independently associated with a higher likelihood of myocardial infarction.^25^

### Future directions

Future studies should be focused not only on the data from a single blood pressure reading but also on blood pressure variability through hospital stay while addressing possible factors that can lead to high variability, including appropriate medication reconciliation, as well as strategies to address anxiety, sleep deprivation, psychological stress, and pain that can also elevate blood pressure and trigger PRN BP medication and contribute to BP variability.

### Strengths and limitation

The findings from the current study should be interpreted within the context of its limitations and regarded as hypothesis-generating. First, all included studies were observational despite best efforts to reduce confounding with propensity score match; the individual patient risk for stroke and AKI were not taken into account. Second, there were mild variations in the definition of intensified and non-intensified blood pressure across the studies included, despite the same principle in all studies: patients received more anti-hypertensive medication in the intensified group compared to the conservative management group. Our analysis is limited by significant heterogeneity with a potential contributor such as multiple classes of anti-hypertensive medications and different proportions of this medication in each study. Potential confounders may be the more frequent IV administration route and more frequent hydralazine use in the intensified BP control group. These factors highlight the need for further randomized studies to answer these clinical questions. All studies included only older adults, with a mean age of 69.9 years in the intensified management arm and 70.1 years in the conservative arm; thus, our findings are not generalizable to younger populations.

### Conclusion

In this meta-analysis of patients with noncardiac admissions, our findings suggest that intensified blood pressure control during admission and on discharge was associated with a significantly higher chance of stroke, AKI, and prolonged length of stay. The increased chance of MI was not statistically significant. These findings underscore the potential harm of intensified blood pressure control and emphasize the need for further RCT.

## Supporting information

Data extraction sheet ( for systematic review and meta-analysis)

## Data Availability

The authors confirm that the data supporting the findings of this study are available within the article [and/or] its supplementary materials.

## Abbreviations

PRISMA: Preferred reporting items for systematic reviews and meta-analysis
PROSPERO: International Prospective Register of Systematic Reviews
RCT: Randomized controlled trial
ROBINS – I: Risk of Bias in Non-randomized Studies of interventions
PRN: Pro re nata (as needed)
AKI: Acute Kidney Injury
CKD: Chronic kidney disease

---

**Supplementary Table 1.** Critical appraisal of individual studies according to the Cochrane Collaboration’s tool for assessing risk of bias in non-randomized trials. **Risk of bias assessment, ROBINS-I**

| Study | Bias due to confounding | Bias in selection of participants | Bias in classification of interventions | Bias due to deviations from intended interventions | Bias due to missing data | Bias in measurement of outcomes | Bias in selection of the reported result | Overall risk of bias judgment |
| --- | --- | --- | --- | --- | --- | --- | --- | --- |
| Anderson | Moderate | Low | Low | Low | Low | Low | Low | Low |
| Radhika | Moderate | Low | Low | Low | Low | Low | Low | Low |
| Mohandas | Moderate | Low | Low | Low | Low | Low | Low | Low |
| Canales | Moderate | Low | Low | Low | Low | Low | Low | Low |

**Supplementary Figure 1.**
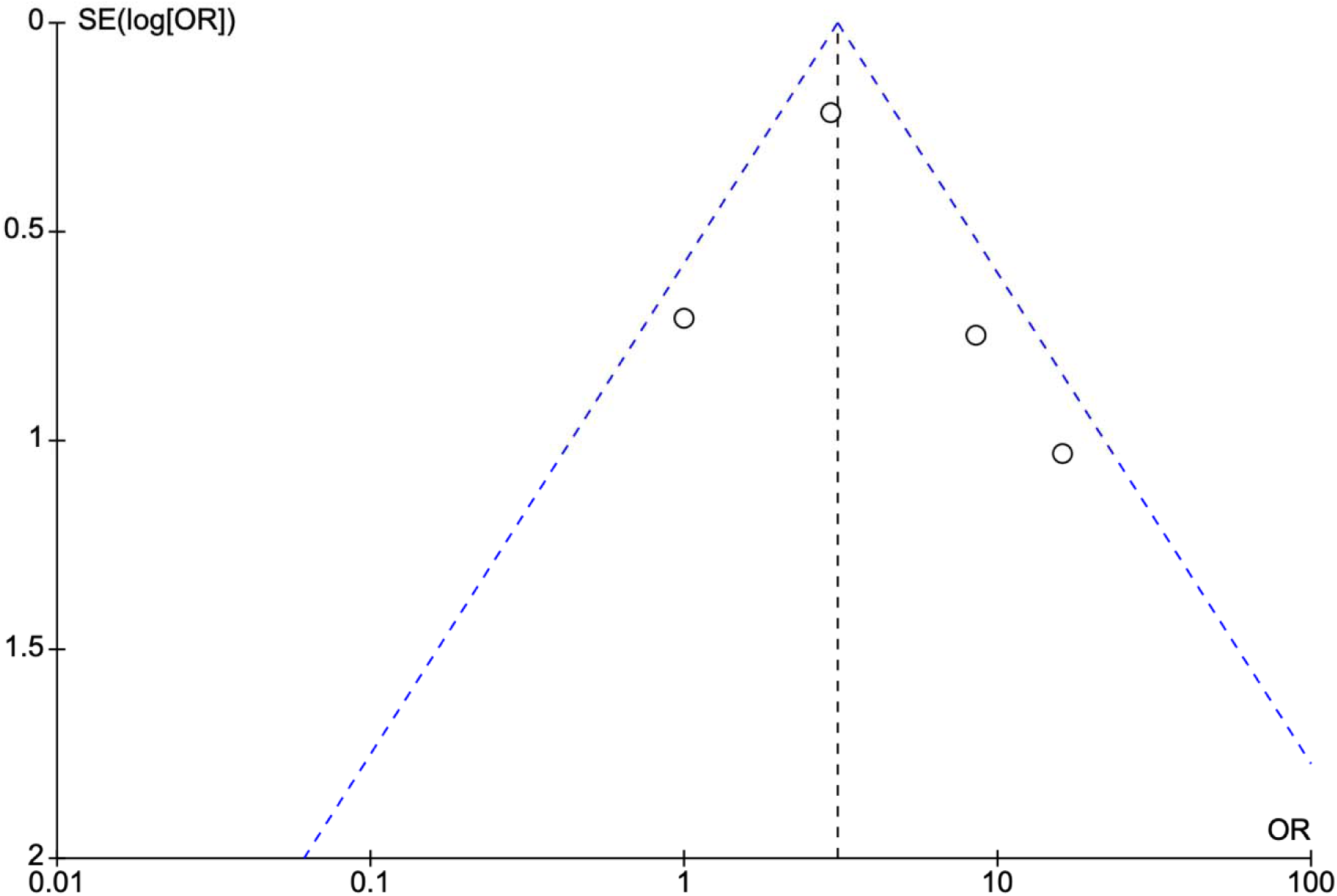
Funnel plot for risk of stroke in intensified and non-intensified blood pressure control showed no definitive evidence of publication bias.

